# High-cited favorable studies for COVID-19 treatments ineffective in large trials

**DOI:** 10.1101/2022.01.11.22269097

**Authors:** John P.A. Ioannidis

## Abstract

**Importance:** COVID-19 has resulted in massive production, publication and wide dissemination of clinical studies trying to identify effective treatments. However, several widely touted treatments failed to show effectiveness in large well-done randomized controlled trials (RCTs).

**Objective:** To evaluate for COVID-19 treatments that showed no benefits in subsequent large RCTs how many of their most-cited clinical studies had declared favorable results for these interventions.

**Methods:** Scopus (last update December 23, 2021) identified articles on lopinavir-ritonavir, hydroxycholoroquine/azithromycin, remdesivir, convalescent plasma, colchicine or interferon (index interventions) that represented clinical trials and that had received >150 citations. Their conclusions were assessed and correlated with study design features. The ten most recent citations for the most-cited article on each index intervention were examined on whether they were critical to the highly-cited study. Altmetric scores were also obtained.

**Findings:** 40 articles of clinical studies on these index interventions had received >150 citations (7 exceeded 1,000 citations). 20/40 (50%) had favorable conclusions and 4 were equivocal. Highly-cited articles with favorable conclusions were rarely RCTs while those without favorable conclusions were mostly RCTs (3/20 vs 15/20, p=0.0003). Only 1 RCT with favorable conclusions had sample size >160. Citation counts correlated strongly with Altmetric scores, in particular news items. Only 9 (15%) of 60 recent citations to the most highly-cited studies with favorable or equivocal conclusions were critical to the highly-cited study.

**Conclusion:** Many clinical studies with favorable conclusions for largely ineffective COVID-19 treatments are uncritically heavily cited and disseminated. Early observational studies and small randomized trials may cause spurious claims of effectiveness that get perpetuated.

## INTRODUCTION

The search for COVID-19 treatments has ushered in thousands of clinical studies,^1-3^ many promises, several emergency authorizations, and some excellent successes. In particular, large adaptive randomized controlled trials (RCTs) using rapid recruitment of participants from real-world clinical practice were instrumental in documenting benefits with large-scale evidence.^4,5^ By the end of 2021 dexamethasone, tocilizumab, and monoclonal antibody combinations had shown convincing evidence that they reduce mortality in various patient groups and clinical settings.^6-8^ However, the largest randomized controlled trials (RCTs) to-date, RECOVERY^9-13^ and SOLIDARITY,^14^ have also showed no benefit for several other treatments. These treatments include lopinavir/ritonavir,^9^ hydroxycholoroquine,^10,14^ azithromycin,^11^ remdesivir,^14^ convalescent plasma,^12^ colchicine^13^ and interferon.^14^ All these interventions with disappointing results in the large trials had been touted as being highly promising and effective in earlier, mostly smaller studies. Each of these treatments have been debated heavily in both scientific and lay circles, often vehemently so. The unfavorable results of the large trials have also led in several situations to reversals of emergency authorizations and/or removal of these interventions from the list of recommended treatments in guidelines.

The ability of the scientific literature to move forward and adopt the newer, more sober evidence is unknown. Important questions can be asked. Has the emergence of the unfavorable results from the large, carefully conducted RCTs managed to change the pervasive presence of these treatments in the scientific literature? Do authors continue to cite the highly promising early clinical studies with the favorable results? If so, are these citations critical of the original promising studies and do they cite also the well-powered and carefully executed RCTs that had null results?

The present analysis aimed to evaluate how many of the most-cited clinical studies in the literature have been favorable for interventions that failed to show a survival benefit for COVID-19 patients in the two largest, well-powered RCTs, RECOVERY^9-13^ and SOLIDARITY.^14^ The analysis also aimed to evaluate whether study design features are associated with favorable conclusions; how citations have tracked against media and social media interest (as captured by the Altmetric score); and whether citing articles to the most-cited studies were critical and whether they cited also the refuting large RCTs.

## METHODS

### Eligible highly cited studies and search strategy

A Scopus search (last update 12/23/2021) sought articles published in 2020-2021 with lopinavir-ritonavir, hydroxycholoroquine, azithromycin, remdesivir, convalescent plasma, colchicine or interferon (title-abstract-keywords). Articles were eligible if they were clinical studies that pertained to any of these index interventions and that had received over 150 citations in Scopus by the search date. The citation threshold was pre-specified but it is arbitrary. For articles published very recently, >150 citations is extremely rare (<1% of total). Therefore, these articles are exceptionally influential in the scientific literature.

The index treatments were selected because they have been evaluated in the two largest RCTs of therapeutic interventions for COVID-19 and have shown no significant benefit for the primary outcome of mortality and no other signals of any substantive benefit for other clinically important outcomes. The relative risks for the mortality outcome in RECOVERY was 1.03 (95% confidence interval [CI] 0.91-1.17) based on 5040 randomized participants for lopinavir-ritonavir,^9^ 1.09 (95% CI, 0.97-1.23) based on 4716 randomized participants for hydroxychloroquine,^10^ 0.97 (95% CI, 0.87-1.07) based on 7763 randomized participants for azithromycin,^11^ 1.00 (95% CI, 0.93-1.07) based on 11558 randomized participants for convalescent plasma,^12^ and 1.01 (95% CI 0.93-1.10) based on 11340 randomized participants for colchicine.^13^ The relative risks for the mortality outcome in SOLIDARITY,^14^ a trial sponsored by the World Health Organization to assess repurposed drugs for COVID-19,^13^ were 0.95 (95% CI, 0.81-1.11) based on 5451 randomized participants for remdesivir, 1.19, 95% CI, 0.89-1.59) based 1853 randomized participants for remdesivr, 1.00 (95% CI, 0.79-.125) based on 2771 randomized participants for lopinavir, and 1.16 (95% CI, 0.96-1.39) based on 4100 randomized participants for interferon.

Among the highly-cited articles retrieved on these index treatments, any study design was eligible (RCT, non-randomized controlled, uncontrolled study [including also case reports and case series]). Clinical studies where the index intervention(s) was involved along with other treatment(s) were also eligible, unless the clinical study focused on the other treatment(s) (e.g. tested a new treatment versus standard of care and used an index intervention as a common backbone for both arms) or found that a new treatment is superior to an index intervention (in which case, one cannot conclude whether the index intervention by itself is effective or not). Retracted articles and their retraction notices were also excluded.

### Data extraction

Data extraction on the eligible highly-cited studies recorded the evaluated interventions, the design (randomized or not), sample size, and number of deaths. The conclusions of the authors of each eligible highly-cited study were categorized as favorable (claiming a benefit and/or safety without harm for an index treatment); unfavorable (claiming no benefit and/or a safety problem for an index treatment); or equivocal when there was a mixed message or potential benefit seen in some particular analysis or endpoint.

### Media and social media impact

Media and social media impact was assessed by the Altmetric score. Information was also obtained through Altmetric on the rank of the paper across all scientific papers tracked by Altmetric and on the number of news items and tweets mentioning each highly-cited paper. Correlations between citation counts and Altmetric score, news items, and tweets were estimated with Pearson coefficients.

### Citation content analysis

For each index treatment, the 10 most recent citing papers to its most highly-cited study with favorable or equivocal results were probed to assess whether the citations were critical to the highly-cited study and to identify whether the citing papers also cited the large trials with unfavorable results (RECOVERY and/or SOLIDARITY)^9-14^ – either the final peer-reviewed publications, or the preprints, or at least some press release or other mention of these unfavorable results. Unfavorable results were announced by RECOVERY for hydroxychloroquine on June 5, 2020, for lopinavir-ritonavir on June 29, 2020, for azithromycin of December 14, 2020, for convalescent plasma on January 15, 2020, and for colchicine on March 5, 2021 and by SOLIDARITY on hydroxychloroquine, remdesivir, lopinavir, and interferon on October 15, 2020. Therefore, it is very likely that there was sufficient time for the authors of the examined citing papers to be aware of these results when they wrote or at least when they revised their papers. The most recent citing papers as of December 27, 2021 were retrieved from Scopus.

## RESULTS

### Highly-cited clinical studies on COVID-19 index treatments

The Scopus search yielded 63803 results, of which 465 published items were highly-cited with >150 citations. 45 of the 465 pertained to any of the index interventions being used in clinical studies. A retracted paper and its retraction notice were excluded, and another 3 were excluded because the favored treatment (baricitinib, arbidol) was not an index intervention. The 40 eligible articles are shown in Table 1.^9,10,14-51^

**Table 1.**
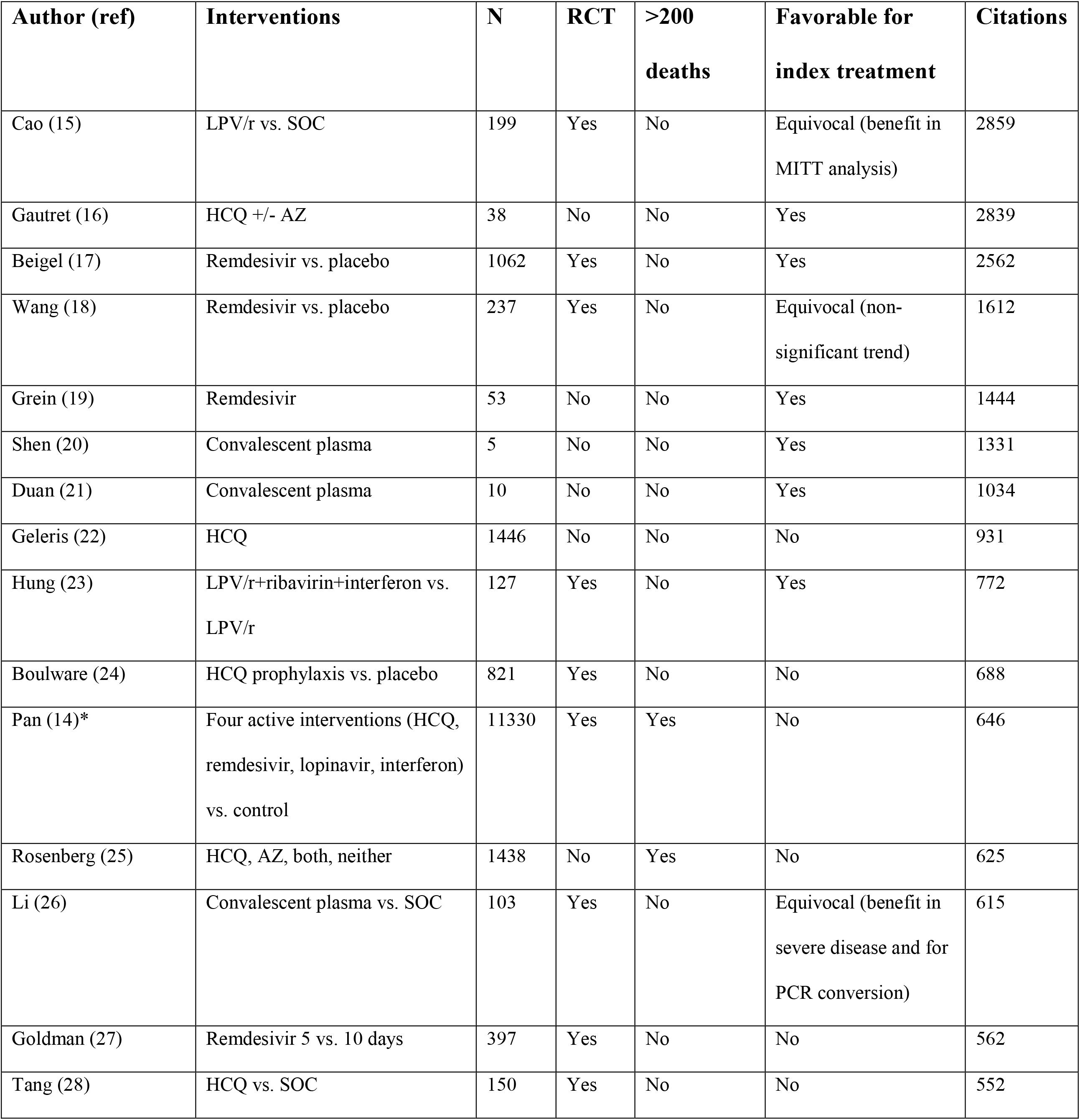
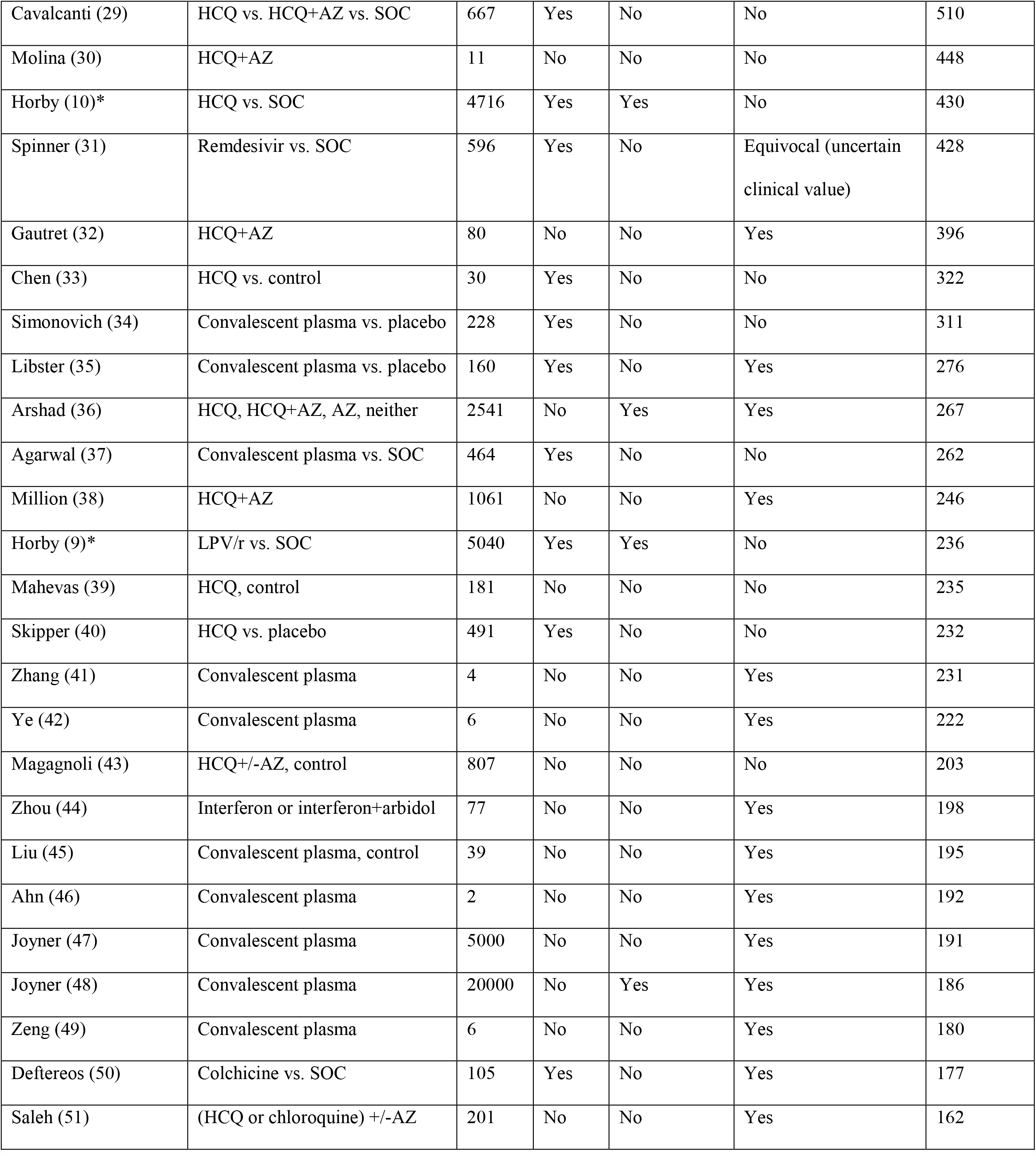
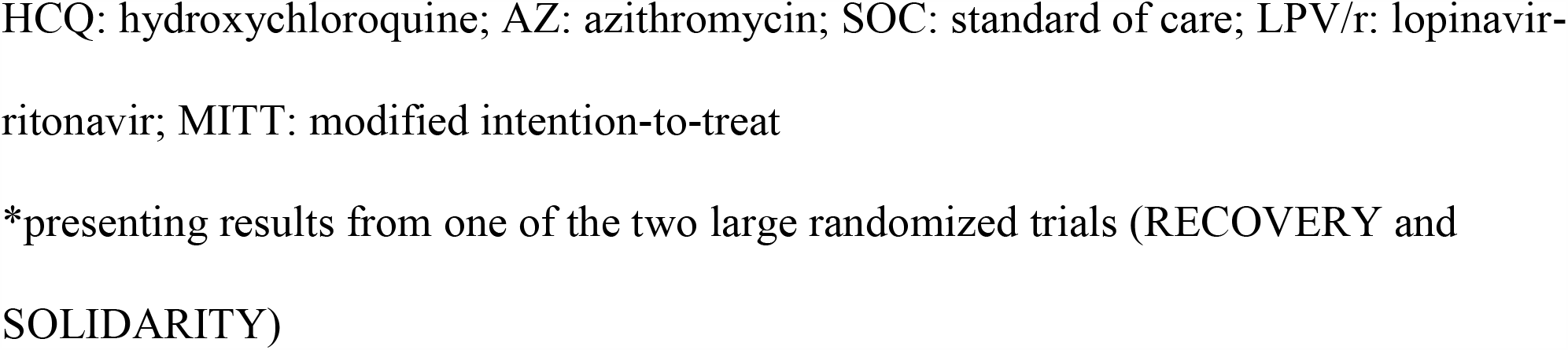
Clinical studies with more than 150 Scopus citations that assess COVID-19 treatments that have shown no benefit in large trials (RECOVERY and SOLIDARITY).

| **Author (ref)** | **Interventions** | **N** | **RCT** | **>200 deaths** | **Favorable for index treatment** | **Citations** |
| --- | --- | --- | --- | --- | --- | --- |
| Cao (15) | LPV/r vs. SOC | 199 | Yes | No | Equivocal (benefit in MITT analysis) | 2859 |
| Gautret (16) | HCQ +/- AZ | 38 | No | No | Yes | 2839 |
| Beigel (17) | Remdesivir vs. placebo | 1062 | Yes | No | Yes | 2562 |
| Wang (18) | Remdesivir vs. placebo | 237 | Yes | No | Equivocal (non-significant trend) | 1612 |
| Grein (19) | Remdesivir | 53 | No | No | Yes | 1444 |
| Shen (20) | Convalescent plasma | 5 | No | No | Yes | 1331 |
| Duan (21) | Convalescent plasma | 10 | No | No | Yes | 1034 |
| Geleris (22) | HCQ | 1446 | No | No | No | 931 |
| Hung (23) | LPV/r+ribavirin+interferon vs. LPV/r | 127 | Yes | No | Yes | 772 |
| Boulware (24) | HCQ prophylaxis vs. placebo | 821 | Yes | No | No | 688 |
| Pan (14)* | Four active interventions (HCQ, remdesivir, lopinavir, interferon) vs. control | 11330 | Yes | Yes | No | 646 |
| Rosenberg (25) | HCQ, AZ, both, neither | 1438 | No | Yes | No | 625 |
| Li (26) | Convalescent plasma vs. SOC | 103 | Yes | No | Equivocal (benefit in severe disease and for PCR conversion) | 615 |
| Goldman (27) | Remdesivir 5 vs. 10 days | 397 | Yes | No | No | 562 |
| Tang (28) | HCQ vs. SOC | 150 | Yes | No | No | 552 |
| Cavalcanti (29) | HCQ vs. HCQ+AZ vs. SOC | 667 | Yes | No | No | 510 |
| Molina (30) | HCQ+AZ | 11 | No | No | No | 448 |
| Horby (10)* | HCQ vs. SOC | 4716 | Yes | Yes | No | 430 |
| Spinner (31) | Remdesivir vs. SOC | 596 | Yes | No | Equivocal (uncertain clinical value) | 428 |
| Gautret (32) | HCQ+AZ | 80 | No | No | Yes | 396 |
| Chen (33) | HCQ vs. control | 30 | Yes | No | No | 322 |
| Simonovich (34) | Convalescent plasma vs. placebo | 228 | Yes | No | No | 311 |
| Libster (35) | Convalescent plasma vs. placebo | 160 | Yes | No | Yes | 276 |
| Arshad (36) | HCQ, HCQ+AZ, AZ, neither | 2541 | No | Yes | Yes | 267 |
| Agarwal (37) | Convalescent plasma vs. SOC | 464 | Yes | No | No | 262 |
| Million (38) | HCQ+AZ | 1061 | No | No | Yes | 246 |
| Horby (9)* | LPV/r vs. SOC | 5040 | Yes | Yes | No | 236 |
| Mahevas (39) | HCQ, control | 181 | No | No | No | 235 |
| Skipper (40) | HCQ vs. placebo | 491 | Yes | No | No | 232 |
| Zhang (41) | Convalescent plasma | 4 | No | No | Yes | 231 |
| Ye (42) | Convalescent plasma | 6 | No | No | Yes | 222 |
| Magagnoli (43) | HCQ+/-AZ, control | 807 | No | No | No | 203 |
| Zhou (44) | Interferon or interferon+arbidol | 77 | No | No | Yes | 198 |
| Liu (45) | Convalescent plasma, control | 39 | No | No | Yes | 195 |
| Ahn (46) | Convalescent plasma | 2 | No | No | Yes | 192 |
| Joyner (47) | Convalescent plasma | 5000 | No | No | Yes | 191 |
| Joyner (48) | Convalescent plasma | 20000 | No | Yes | Yes | 186 |
| Zeng (49) | Convalescent plasma | 6 | No | No | Yes | 180 |
| Deftereos (50) | Colchicine vs. SOC | 105 | Yes | No | Yes | 177 |
| Saleh (51) | (HCQ or chloroquine) +/-AZ | 201 | No | No | Yes | 162 |

HCQ: hydroxychloroquine; AZ: azithromycin; SOC: standard of care; LPV/r: lopinavir-ritonavir; MITT: modified intention-to-treat
\*presenting results from one of the two large randomized trials (RECOVERY and SOLIDARITY)

### Favorable conclusions

Of 40 eligible studies, 20 (50%) had favorable conclusions for the index treatments, 4 were equivocal and 16 were unfavorable. The unfavorable group included the publication of SOLIDARITY trial itself^14^ and the publications of the hydroxychloroquine^10^ and lopinavir-ritonavir^9^ assessments from the RECOVERY trial. Of 7 articles that exceeded 1,000 citations, 5 had favorable conclusions, and 2 were equivocal (one described significant benefit in modified intention-to-treat analysis, and the other mentioned a non-significant trend for clinical improvement).

### Correlates of favorable conclusions

The highly-cited articles with favorable conclusions were far less likely to be randomized controlled trials than the other highly-cited articles (3/20 versus 15/20, exact p=0.0003). The few randomized controlled trials with favorable conclusions tended to be smaller than the others (median sample size 160 versus 464). Of the 6 studies with at least 200 deaths, all three randomized trials did not reach favorable conclusions; 2 of 3 non-randomized studies did.

### Altmetric scores

As shown in Supplementary Table 1, all 40 highly-cited papers had very high Altmetric scores placing them at the top 5% of published papers and 24/40 had extraordinarily high Altmetric scores placing them among the 2000 highest Altmetric scored papers of all science of all times. Five papers were among the top-200 of all science of all times. There was a strong correlation between the Altmetric score and the number of citations (r=0.74) (Figure 1). The correlation of the number of citations was stronger with the number of news items (r=0.81) and more modest with the number of tweets (n=0.47).

**Figure 1.**
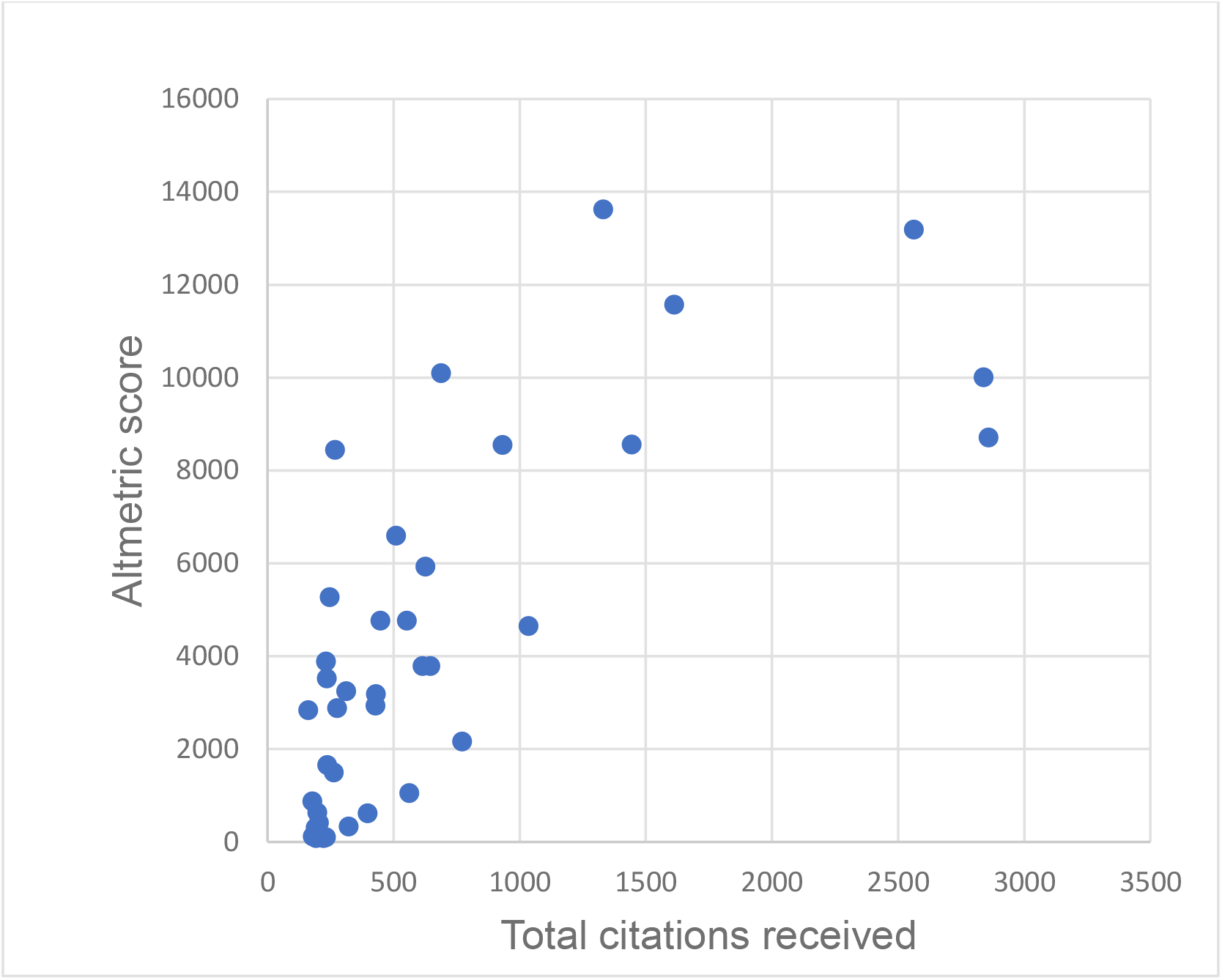
Correlation between citation counts and Altmetric scores for the eligible highly-cited papers

Favorable papers did not have higher media and social media mentions than other papers. For example, Altmetric values in the top-2000 of all science occurred in 9/20 favorable, 4/4 equivocal and 11/16 unfavorable papers (exact p=0.10 for the comparison of papers with favorable conclusions versus other papers).

### Analysis of most recent citing articles

Only 9 (15%) of 60 recent citations to the most highly-cited studies with favorable or equivocal conclusions were critical to the highly-cited study (Table 2). Citing papers uncommonly (8/60, 13%) cited the respective RECOVERY or SOLIDARITY results.

**Table 2.**
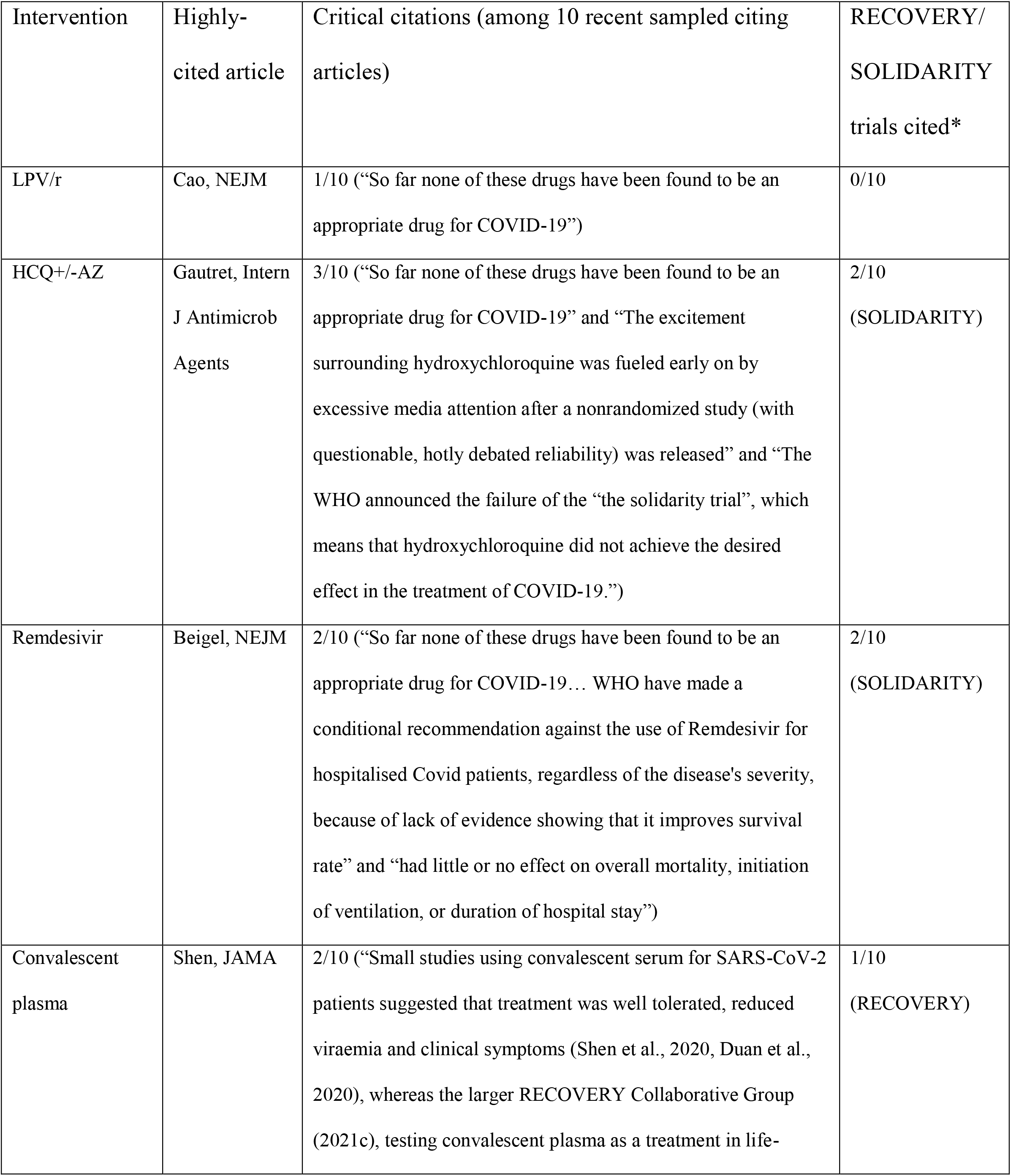
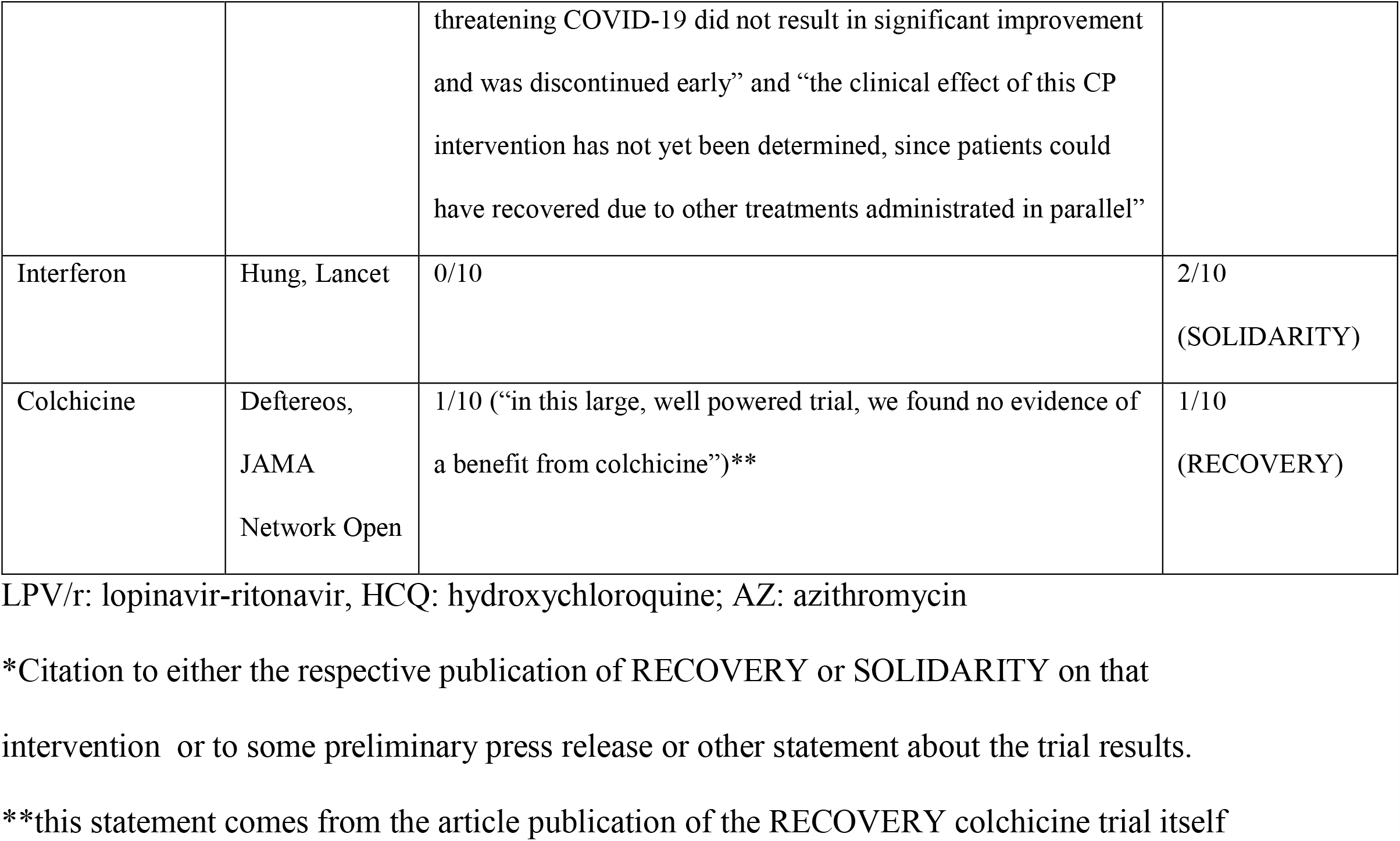
Qualitative analysis of recent citations to the most highly-cited article for each index treatment that reached favorable or equivocal conclusions.

## DISCUSSION

This analysis demonstrates that many highly-cited clinical studies favor COVID-19 treatments that have shown no benefits in large, well-powered randomized trials. Most favorable studies are not randomized or are even uncontrolled, but they can still exercise strong, persistent influence on the scientific literature. Citation counts track well with strong presence of these studies in media and social media. The most highly-cited studies on these interventions have either entirely or partially favorable conclusions and the citations that they continue to receive are rarely critical of them.

Citations are a measure of the influence of a research paper across the broad scientific literature. Various manifestations of citation bias have been demonstrated in other clinical and scientific fields well before the COVID-19 era.^52-57^ In principle, studies with “positive” results tend to be more heavily cited than studies with “negative” results on the same topic. The citation bias creates a distorted picture for the perception of the scientific literature at large. Repeated mention of the most favorable results gives the allusion that they are more likely to represent the truth, while this may not be the case. The COVID-19 scientific literature is also unique in terms of the massive volume of papers produced^58,59^ (and thus also citations generated) within a very limited timeframe. Very few studies in the history of medicine have ever received the number of citations received by the most highly-cited COVID-19-related papers – including those whose promises could not be validated by large, well-powered RCTs.

The highly polarized and charged situation surrounding the COVID-19 pandemic may have further complicated matters and intensified the bias. Several of these treatments have received tremendous attention not only in the scientific literature, but also in the wider society and they have both strong supporters and strong critics. Many of the highly-cited papers analyzed here have also reached astronomical Altmetric scores, due to their massive discussion in media and social media. Altmetric scores correlated well with citation counts. The correlation was more prominent when news items were considered, while tweets had a more modest correlation with citation count. Altmetric score analyses have shown^60^ that media and social media attention may remain high even for fully retracted papers.

Some caveats need to be acknowledged. The most important limitation of this analysis is that the large trials may not necessarily be a perfect gold standard. No single clinical study can claim to possess the perfect truth, no matter how well it is conducted and how well it is protected from bias. The CIs of the large trials cannot exclude very small benefits on survival – or small harms. These trials have also shown no benefit also on other outcomes. However, small benefits (or harms) for these outcomes are also not possible to exclude with perfect certainty. Moreover, beneficial effects for some treatments may still exist in circumscribed, special circumstances, with different dosing regimens, and in specific patient subgroups that may have been outside the eligibility criteria of the large RCTs or may have been under-represented in these large RCTs. However, similar concerns and speculative counter-arguments may be raised almost in any clinical topic, especially by those who still believe that a treatment may have merits despite its poor performance in very large trials.^61^

It should also be acknowledged that not all guidelines have removed these treatments from the list of interventions that they recommend. Remdesivir is probably the most notable example in this regard. It is not recommended by the European Respiratory Society^62^ and the World Health Organization has issued a conditional recommendation *against* its use.^63^ Conversely, the US National Institutes of Health (NIH) list remdesivir very prominently among the very few treatments that they recommend.^64^ Subjective interpretation of the evidence is obvious in these differing opinions. Moreover, there may be overlap or connection between the researchers and institutions who perform the clinical studies and those that issue the recommendations and guidelines. For example, the most-cited favorable trial on remdesivir was spearheaded by NIH.^17^ Moreover, a consequence of trusting a treatment as being effective is that it becomes attractive, if not necessary, to use it as background treatment or as a comparator when a new intervention is to be evaluated. In the case of remdesivir, NIH-spearheaded trials have already done this. E.g. they have compared interferon+remdesivir versus remdesivir alone and claimed to find no benefit for interferon;^65^ and baricitinib+remdesivir versus remdesivir alone and claimed to find a benefit for the addition of baricitinib.^66^ Retrospectively, these study designs are poor choices if remdesivir is indeed ineffective. They would be even highly misleading if remdesivir happens to be harmful.

Acknowledging these caveats and some residual uncertainty, it is more likely that these treatments overall don’t save lives eventually. The most recent meta-analyses are also consistent with this interpretation.^67-71^ For hydroxychloroquine, a recent collaborative meta-analysis even shows nominally statistically significantly increased mortality.^72^

Prior experience from the in-depth analysis of persistent high citations of non-validated papers in other fields may offer us insights on why this situation arose also for COVID-19 treatments. One empirical evaluation^61^ assessed in-depth the citation patterns of extremely highly-cited papers on the benefits of beta carotene for cancer prevention, on estrogens for Alzheimer’s dementia and on alpha tocopherol for cardiovascular disease. Despite the emergence of large RCTs with unfavorable results for these interventions, the observational studies that made the original promises continued to be heavily cited long after the “negative” RCT results had been published. Their citations were either ignoring the refuting RCTs (for beta carotene), or raising numerous counterarguments against them (for estrogen and alpha tocopherol). Similarly, in psychology, where several major claims had been found to be irreproducible in pre-registered reproducibility assessments done by many teams, frequent citation of the original claims has continued unperturbed after their non-reproduction.^73^ The citing articles uncommonly take a critical stance against the original claims.^73^ In other areas where evidence is heavily centered on biological considerations, citation networks citing some preferred papers may create a perpetuated distortion of what is the established knowledge. This has been seen in empirical evaluations in genetics of amygdala activation^74^ and in pathology of inclusion body myositis.^75^ Another empirical evaluation even found evidence of so-called “affirmative citation bias”.^52^ The citations to the critical articles that had thoroughly debunked prior beliefs were mostly affirmative, in favor of the original beliefs that had been debunked. The authors concluded that even criticism itself may paradoxically reinforce the establishment of debunked prior beliefs.^52^

These observations suggest that often science is organized in cliques or schools of thought that are recalcitrant to the provision and acceptance of contrarian evidence. This may have happened also in the case of the early promising COVID-19 treatments. Moreover, in COVID-19, given the vast attention devoted to the topic, the citation rates of these non-validated treatment benefits are even more extraordinary. Furthermore, the overall impact of their dissemination reverberates across wider societal circles, not just scientific groups. The advent of very large RCTs did not suffice to perturb much this intense dissemination.

In conclusion, one should avoid putting much trust to highly promising results from early observational studies and small randomized trials of new or repurposed treatments. For serious diseases, like COVID-19, evidence on mortality endpoints should be sought. Pilot studies should not be abandoned or dejected, and they do have some value in offering early insights. However, they should be seen with great caution and with tempered enthusiasm. Large trials with flexible designs that allow obtaining large-scale rigorous evidence in a timely manner have been a major success during the pandemic^3-5^ and their use should be promoted further.

## Data Availability

All data produced in the present work are contained in the manuscript

---

**Supplementary Table 1.**
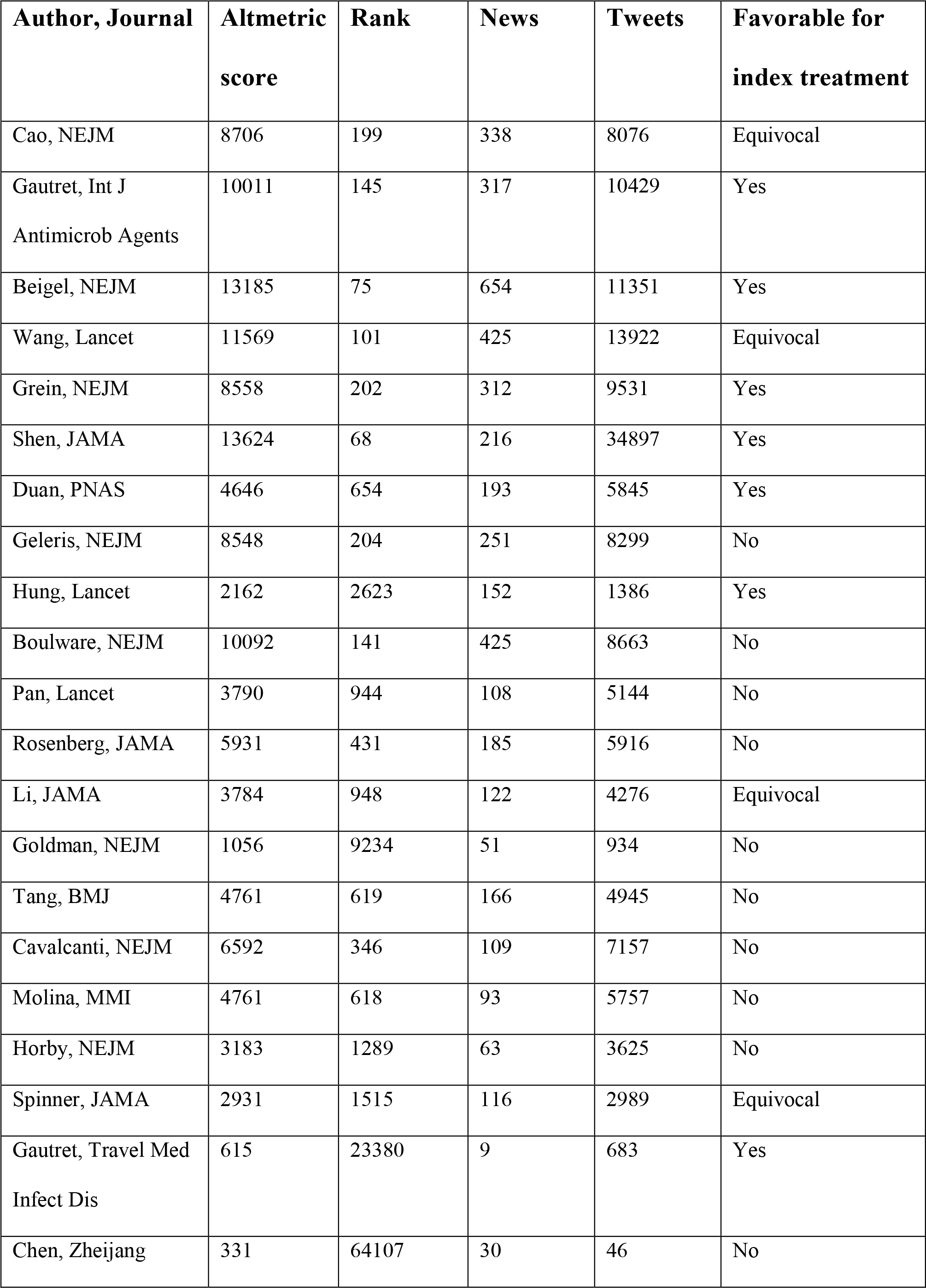
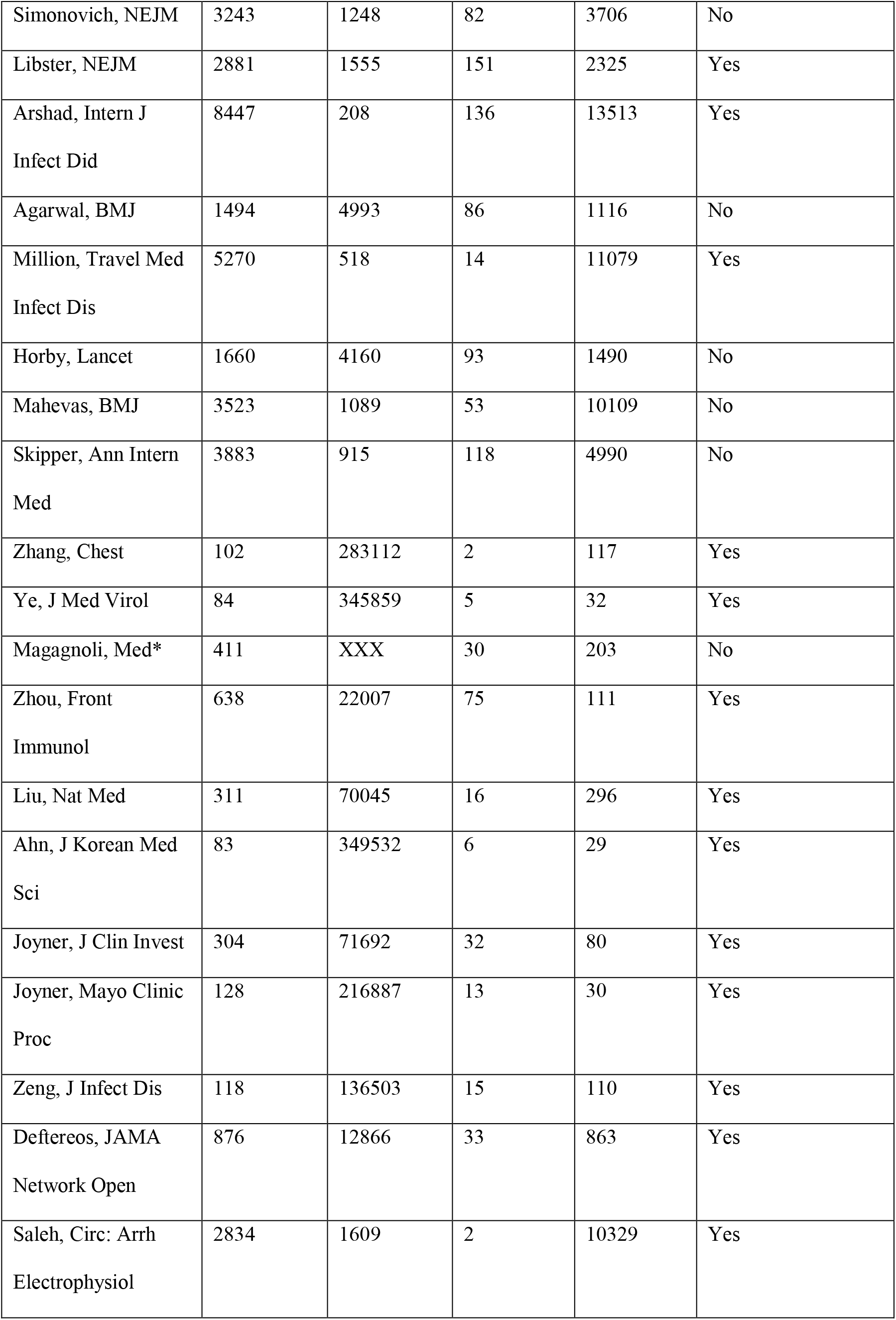
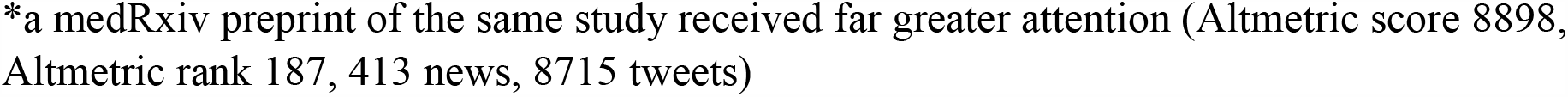
Altmetric scores for the highly cited clinical studies

| **Author, Journal** | **Altmetric score** | **Rank** | **News** | **Tweets** | **Favorable for index treatment** |
| --- | --- | --- | --- | --- | --- |
| Cao, NEJM | 8706 | 199 | 338 | 8076 | Equivocal |
| Gautret, Int J Antimicrob Agents | 10011 | 145 | 317 | 10429 | Yes |
| Beigel, NEJM | 13185 | 75 | 654 | 11351 | Yes |
| Wang, Lancet | 11569 | 101 | 425 | 13922 | Equivocal |
| Grein, NEJM | 8558 | 202 | 312 | 9531 | Yes |
| Shen, JAMA | 13624 | 68 | 216 | 34897 | Yes |
| Duan, PNAS | 4646 | 654 | 193 | 5845 | Yes |
| Geleris, NEJM | 8548 | 204 | 251 | 8299 | No |
| Hung, Lancet | 2162 | 2623 | 152 | 1386 | Yes |
| Boulware, NEJM | 10092 | 141 | 425 | 8663 | No |
| Pan, Lancet | 3790 | 944 | 108 | 5144 | No |
| Rosenberg, JAMA | 5931 | 431 | 185 | 5916 | No |
| Li, JAMA | 3784 | 948 | 122 | 4276 | Equivocal |
| Goldman, NEJM | 1056 | 9234 | 51 | 934 | No |
| Tang, BMJ | 4761 | 619 | 166 | 4945 | No |
| Cavalcanti, NEJM | 6592 | 346 | 109 | 7157 | No |
| Molina, MMI | 4761 | 618 | 93 | 5757 | No |
| Horby, NEJM | 3183 | 1289 | 63 | 3625 | No |
| Spinner, JAMA | 2931 | 1515 | 116 | 2989 | Equivocal |
| Gautret, Travel Med Infect Dis | 615 | 23380 | 9 | 683 | Yes |
| Chen, Zheijang | 331 | 64107 | 30 | 46 | No |
| Simonovich, NEJM | 3243 | 1248 | 82 | 3706 | No |
| Libster, NEJM | 2881 | 1555 | 151 | 2325 | Yes |
| Arshad, Intern J<br>Infect Dis | 8447 | 208 | 136 | 13513 | Yes |
| Agarwal, BMJ | 1494 | 4993 | 86 | 1116 | No |
| Million, Travel Med<br>Infect Dis | 5270 | 518 | 14 | 11079 | Yes |
| Horby, Lancet | 1660 | 4160 | 93 | 1490 | No |
| Mahevas, BMJ | 3523 | 1089 | 53 | 10109 | No |
| Skipper, Ann Intern<br>Med | 3883 | 915 | 118 | 4990 | No |
| Zhang, Chest | 102 | 283112 | 2 | 117 | Yes |
| Ye, J Med Virol | 84 | 345859 | 5 | 32 | Yes |
| Magagnoli, Med* | 411 | XXX | 30 | 203 | No |
| Zhou, Front<br>Immunol | 638 | 22007 | 75 | 111 | Yes |
| Liu, Nat Med | 311 | 70045 | 16 | 296 | Yes |
| Ahn, J Korean Med<br>Sci | 83 | 349532 | 6 | 29 | Yes |
| Joyner, J Clin Invest | 304 | 71692 | 32 | 80 | Yes |
| Joyner, Mayo Clinic<br>Proc | 128 | 216887 | 13 | 30 | Yes |
| Zeng, J Infect Dis | 118 | 136503 | 15 | 110 | Yes |
| Deftereos, JAMA<br>Network Open | 876 | 12866 | 33 | 863 | Yes |
| Saleh, Circ: Arrh<br>Electrophysiol | 2834 | 1609 | 2 | 10329 | Yes |

\*a medRxiv preprint of the same study received far greater attention (Altmetric score 8898, Altmetric rank 187, 413 news, 8715 tweets)

## Data sharing

all data are in the paper and in the supplementary table 1.

## Patient involvement

none

